# Heterozygous Nonsense Variants in the Ferritin Heavy Chain Gene *FTH1* Cause a Novel Pediatric Neuroferritinopathy

**DOI:** 10.1101/2023.01.30.23285099

**Authors:** Joseph T Shieh, Jesus A Tintos-Hernández, Chaya N. Murali, Monica Penon-Portmann, Marco Flores-Mendez, Adrian Santana, Joshua A. Bulos, Kang Du, Lucie Dupuis, Nadirah Damseh, Roberto Mendoza-Londoño, Camilla Berera, Julieann C Lee, Joanna J Phillips, César A P F Alves, Ivan J Dmochowski, Xilma R Ortiz-González

**Author notes:** These authors contributed equally to this work (Joseph T Shieh, Jesus A Tintos-Hernández).

## Abstract

Ferritin, the iron storage protein, is composed of light and heavy chain subunits, encoded by *FTL* and *FTH1*, respectively. Heterozygous variants in *FTL* cause hereditary neuroferritinopathy, a type of neurodegeneration with brain iron accumulation (NBIA). Variants in *FTH1* have not been previously associated with neurologic disease. We describe the clinical, neuroimaging, and neuropathology findings of five unrelated pediatric patients with *de novo* heterozygous *FTH1* variants. Children presented with developmental delay, epilepsy, and progressive neurologic decline. Nonsense *FTH1* variants were identified using whole exome sequencing, with a recurrent *de novo* variant (p.F171*) identified in three unrelated individuals. Neuroimaging revealed diffuse volume loss, features of pontocerebellar hypoplasia and iron accumulation in the basal ganglia. Neuropathology demonstrated widespread ferritin inclusions in the brain. Patient-derived fibroblasts were assayed for ferritin expression, susceptibility to iron accumulation, and oxidative stress. Variant *FTH1* mRNA transcripts escape nonsense-mediated decay (NMD), and fibroblasts show elevated ferritin protein levels, markers of oxidative stress, and increased susceptibility to iron accumulation. C-terminus variants in *FTH1* truncate ferritin’s E-helix, altering the four-fold symmetric pores of the heteropolymer and likely diminish iron-storage capacity. *FTH1* pathogenic variants appear to act by a dominant, toxic gain-of-function mechanism. The data support the conclusion that truncating variants in the last exon of *FTH1* cause a novel disorder in the spectrum of NBIA. Targeted knock-down of mutant *FTH1* transcript with antisense oligonucleotides rescues cellular phenotypes and suggests a potential therapeutic strategy for this novel pediatric neurodegenerative disorder.

## Introduction

The transition metal iron has essential roles in biological systems and associations with human disease^1^. Given its range of involvement in cellular functions and its potentially toxic characteristics^2^, iron is highly regulated to maintain homeostasis. Ferritin is a ubiquitous, highly conserved, iron-binding protein, which can store up to 4,500 ionized iron atoms. It is the predominant iron storage protein, allowing for iron to be stored intracellularly as Fe^3+^ and released as Fe^2+^ when needed. Ferritin is a heteropolymer composed of light (L) and heavy (H) chains, encoded by the genes *FTL* and *FTH1*, respectively^3^. The *FTH1* gene is the sole gene encoding the ferritin heavy chain. The heavy chain is responsible for ferroxidase activity, and it has also been implicated in iron delivery to the brain^4^. Ferritin accounts for the majority of iron content in the brain, where ferritin H and L chains are distributed heterogeneously: H ferritin predominates in neurons and L ferritin in microglia^5^, with oligodendrocytes containing substantial amounts of both ferritin chains.

Disruption of iron homeostasis in the brain has been linked to neurodegenerative disorders^6,7^ and genetic disorders such as as neurodegeneration with brain iron accumulation (NBIA), which often present with insidious early-onset neurodevelopmental deficits progressing to neurodegenerative features^8,9^. Commonly affected anatomical areas in NBIA are the basal ganglia, particularly the globus pallidum and substantia nigra. The cerebellum and cortex can also be variably involved, depending on the genetic etiology. Curiously, most genes that cause NBIA disorders do not clearly play a role in iron metabolism. The *FTL* gene, encoding the ferritin light chain, is one of the few currently established NBIA genes known to be directly involved in iron homeostasis^10-15^. Recent studies suggest that multiple NBIA disorders are mechanistically linked to a mitochondrial acyl carrier protein^16^, but overall the molecular mechanisms leading to neurodegeneration in those genetic disorders remain incompletely understood.

Heterozygous variants in *FTL* are associated with the autosomal dominant disorder known as hereditary neuroferritinopathy (also known as NBIA3, MIM: 606159)^17,18^. It is characterized by progressive neurologic symptoms and iron accumulation, particularly in the basal ganglia. The most prominent symptoms include chorea and focal dystonia and, less often, parkinsonism^13^. The most common pathogenic variant in *FTL* is c.450insA, which is a founder mutation from the Cumbrian region of northern England, with a mean age of symptom onset at 39 years^13,17^. Neuropathology shows neuronal ferritin aggregates and neuroimaging indicates accumulation in the basal ganglia. Previous studies suggest pathogenic variants associated with *FTL* hereditary neuroferritinopathy map to the C-terminus of the L-ferritin subunit, disrupting the E-helix domain and affecting iron permeability and storage capacity^19-21^.

Despite the ubiquitous nature of the ferritin heavy chain, variants in the *FTH1* gene have not yet been conclusively linked to human disease. A *FTH1* promoter variant has been reported from a single family with type 5 hemochromatosis (MIM: 615517)^22^, but no other *FTH1*-associated disease has been reported to date. Here, we report a novel human NBIA disorder associated with *de novo* nonsense heterozygous variants in the final exon of *FTH1* in five unrelated children with characteristic clinical and imaging findings. The variants escape NMD and lead to ferritin accumulation on neuropathology, neuroimaging findings consistent with neurodegeneration with brain iron accumulation and cellular markers of oxidative stress.

## Material and Methods

### Human Subjects

Patients presented for evaluation to the Children’s Hospital of Philadelphia Neurogenetics clinic, the University of California San Francisco Medical Genetics clinic, Division of Clinical and Metabolic Genetics clinic at The Hospital for Sick Children in Toronto, and Texas Children’s Hospital genetics clinic in Houston, TX. They underwent clinical neuroimaging and exome sequencing as part of the medical evaluation for neurodevelopmental delay and/or progressive neurologic symptoms. Sequencing was performed by CLIA-certified laboratories. Variants in *FTH1* were identified as candidates, considered variants of unknown significance until further information became available. Human Subjects Protocols for the study have received prior approval by the appropriate Institutional Review Boards and informed consent was obtained from subjects. Primary *FTH1*-variant fibroblasts were established from probands 1, 2 and 3 for cellular studies. Control fibroblasts included the unaffected parent of proband 1 (FTH control, i.e. CTRL2) and fibroblasts from Coriell Institute (GM08400, i.e. CTRL1).

### Neuropathology

Autopsy was performed for proband 2. The brain and spinal cord were fixed in 10% buffered formalin and tissues were processed, embedded, and sectioned for histologic analysis, including hematoxylin and eosin staining and Prussian blue iron stain. Immunostaining with anti-ferritin heavy chain antibody FTH1 Abcam (Cat# ab81444) was performed on formalin-fixed, paraffin-embedded tissue sections. An automated immunostainer was used (Discovery Ultra, Ventana Medical Systems, Inc., Tucson, AZ).

### Cell Culture

Primary fibroblast lines were maintained in Dulbecco’s modified Eagle’s medium (DMEM; Life Technologies) supplemented with Glutamax media, 10-15% fetal bovine serum (FBS), 2 mM L-glutamine, 2.5 mM pyruvate, and non-essential amino acids (NEAAs). Fibroblasts were cultured at 37°C under a humidified atmosphere of 5% CO_2_. While not exceeding passage 15, cells with comparable passage numbers were used for all experiments. Fibroblast genotypes were as follows: proband 1 (P1) *FTH1* c.487_490dupGAAT (p.S164*); probands 2 and 3 (P2 and P3) *FTH1* c.512_513delTT p.(F171*); and non-variant controls.

### Immunoblots

Cells were washed in cold PBS and sonicated twice for 10 seconds in cold lysis buffer (30% CHAPS, 120 mM NaCl, 40 mM HEPES pH 7.5, 50 mM NaF, 2 mM NaVO_3_, protease inhibitor cocktail [CST 5871]). Protein concentration was measured with the Bradford reagent (Bio-Rad). Cell lysates (30 μg) were suspended in Laemmli sample buffer (Bio-Rad 161-0737), and proteins were denatured at 95°C for 5 min in the presence of 10% beta-mercaptoethanol and resolved by SDS/PAGE on 12% gels. After completion of electrophoresis, proteins were transferred to PVDF mini transfer stacks (Invitrogen, IB24002), using the iBlot™ 2 Dry Blotting System [Invitrogen, IB21001] with the following blotting parameters: 20 V for 1 min, 23 V for 4 min, 25 V for 2 min. Blots were stained with Ponceau Red to confirm protein transfer. Blots were then saturated with 5% skim milk in PBS containing 0.1% (v/v) Tween 20 (PBS-T) and probed overnight with antibodies directed against FTH (Thermo Fisher Scientific, PA5-19058; 1:1000) or FTL (AB clonal, A1768; 1:1000). Following a wash with PBS-T, the blots were incubated with Near-Infrared (NIR) fluorescent secondary antibodies; IRDye® 680LT Donkey anti-Goat IgG (H + L) and IRDye® 800CW Donkey anti-Rabbit IgG (H + L) [Li-cor, 1:5000 dilution]. Normalization of targets was done by GAPDH (CST, 97166; 1:1000). Immuno-stained bands were detected with the Odyssey NIR Western Blot detection system and quantified by densitometric scanning using the ImageStudio Lite software (Li-Cor Biosciences).

### Gene Expression Studies

Total RNA was extracted with the RNeasy Mini Kit (QIAGEN) and DNase-treated with TURBO DNA-free Set (Thermo Fisher Scientific, AM1907) to remove contaminating DNA from RNA preparations according to manufacturer’s protocol. The concentration and purity of total RNA were assessed with a NanoDrop 8000 spectrophotometer (Thermo Fisher Scientific). mRNAs were then reverse transcribed from 1 μg total RNA with SuperScript IV First-Strand Synthesis System (Thermo Fisher Scientific, 18091050) and random priming according to the manufacturer’s instructions. Quantitative RT-PCR (qRT-PCR) was carried out in a 10μL reaction mixture with a 384 well format using PowerUp™ SYBR™ Green Master Mix (Thermo Fisher Scientific, A25742) according to the manufacturer’s protocol. Specific primer pairs were as follows: TfR primer (forward primer, 5′-CAGCCCAGCAGAAGCATT-3’; reverse primer, 5′-CCAAGAACCGCTTTATCCAG-3’), *FTL* primer (forward primer, 5’-ACCATGAGCTCCCAGATTCGTC-3’; reverse primer, 5’-CACATCATCGCGGTCGAAATAG-3’), *FTH1* primer (forward primer, 5′-TCAAGAAACCAGACTGTGATGA-3’; reverse primer, 5′-AGTTTGTGCAGTTCCAGTAGT-3’), *B2M* (internal control gene) primer (forward primer, 5’-CCAGCGTACTCCAAAGATTCA-3’; reverse primer, 5’-TGGATGAAACCCAGACACATAG-3’). Real-time PCR comprised an initial denaturation at 95°C for 10 min, 40 cycles at 95°C for 15 s and 60°C for 60 s and then a melt curve stage of 95°C for 15 s, 60°C for 60 s and 95°C for 15 s. Procedures were performed in triplicate and data were averaged.

To specifically detect the level of mutant mRNA expressed, a multiplexed locked nucleic acid (LNA) probe-based real-time PCR was designed against the duplication (dupGAAT from proband 1). LNA oligonucleotide monomers are modified with an additional methylene bridge between 2′ oxygen and 4′ carbon of the ribose ring^23^. The modified LNA monomers allowed the development of a short (16-nt) probe, which improved sensitivity and specificity to detect mutated allele expressed transcripts. Oligonucleotides containing LNA were obtained from IDT. The sequences of the primers used are as follows: WT_*FTH1* allele (forward primer, 5’-TGACCACGTGACCAACTT -3’; reverse primer, 5’-CTTAGCTTTCATTATCACTGTCTC; Probe 5’-/YakYel/CAAGCCAGA/ZEN/TTCGGGCGCTCC/3IABkFQ/3’). Proband 1 *FTH* c.487_490dupGAAT (5’-TACCTGAATGAGCAGGTGAAAG-3’; Reverse 5’-AAGAGATATTCCGCCAAGCC-3’; Probe 5’-/FAM/AGCGCCCG+A+A+TG+A+A+TC/3IABkFQ/-3’), where + represents ribose-modified LNA. Each reaction was normalized utilizing B2M Hs.PT.58v.18759587 (IDT). Multiplexed qRT-PCR reaction was performed at a final volume of 10 μL. Primers were used at the concentration of 400 nM, wild type and mutant probe at 250 nM, each, and 90 ng of cDNA with a 1X PrimeTime Gene Expression Master Mix (IDT 1055771).

### Immunocytochemistry

Immunofluorescence studies were performed using human primary fibroblasts grown on UV-treated glass coverslips in a 24-well plate. Cells were washed with PBS, fixed with 4% paraformaldehyde for 15 min at room temperature (RT) and permeabilized with 0.1% Tween-20 in 10% normal goat serum (Thermo Fisher 50062Z) for 1 h. Primary antibodies: Rabbit anti-FTL (AB Clonal A1768, Goat anti-FTH (Thermo Fisher PA5-19058), mouse anti-LAMP1 (DHSB, H4A3). After incubation with primaries overnight at 4°C, cells were washed 3 times and then incubated with Alexa Fluor® secondary antibodies (Thermo Fisher) for 1 h at RT, then washed 3 times with PBS and mounted on a glass slide using ProLong™ Gold Antifade (Thermo Fisher). Images were acquired with LSM710 (Zeiss) confocal microscope and the maximum intensity projection in the z axis (5 μm) shown.

### Intracellular Iron Quantification

Cellular iron content was assayed by inductively coupled plasma optical emission spectroscopy (ICP-OES), using a Spectro Genesis ICP-OES instrument. Sample preparation was done as follows: To primary fibroblast pellets in a 15 mL centrifuge tube, 200 μL 70% w/w HNO_3_ was added. After thorough mixing on a vortex mixer, the suspension was incubated in a water bath at 50°C for 12 h. After cooling to RT, the resulting solution was added to 4.8 mL deionized water and thoroughly mixed on a vortex mixer to give a sample ready for analysis. In a typical experiment, iron-specific emissions at 238.204, 239.562, 259.941 nm were independently monitored. Signal intensities of the three emission lines for a series of standard solutions containing 0, 5.1994, 19.3728, 50.986, 118.237, 190.891, 502.560, 715.113, 1010.78 ppb iron were used to construct three respective linear calibrations. Reported iron concentrations are averages of values obtained from the three calibrations. Protein content was assayed for each sample and used to normalize iron content values (protein concentration in μg/mL). For all experiments, cells were counted and plated at same density on day 0. For iron treatment, 150 μg/mL of FAC (ferric acid citrate, Sigma) was used for either 3 or 7 days in culture before assaying iron content. To control for different cell density/growth amongst cell lines, an aliquot of all cell pellets was taken prior to iron quantification for protein quantification, which was later used to normalize iron content to protein content per sample.

### Protein Oxidation Assays

An OxyBlot™ Protein Oxidation Detection Kit (Sigma-Aldrich S7150) was used for immunodetection of carbonyl groups, which is a hallmark of the oxidation status of proteins introduced by oxidative reactions, following the manufacturer’s directions. Briefly, two aliquots of each specimen were used. One aliquot was used as a negative control and the other was subjected to conversion to 2,4-dinitrophenylhydrazone (DNP-hydrazone) derivative through reaction with 2,4-dinitrophenylhydrazine (DNPH). Equal amounts (20 μg) of proteins were denatured with SDS (final concentration of 6%) and treated with 1X DNPH solution (20 min) to induce derivatization. Samples designated as negative controls were treated with derivatization-control solution. After incubation, neutralization solution was added to derivatized and negative control samples. Both the treated sample and the negative control were loaded and separated by SDS-PAGE using 12% gels. Sized proteins were transferred, as described above, to a PVDF membrane, non-specific sites were blocked by incubation with blocking/dilution buffer (1% BSA/PBS-T) for 1 h under gentle shaking. Anti-DNP antibody (1:150, Sigma-Aldrich 90451) was added for 1 h at RT. Following 2 washes with PBS-T, blots were incubated with NIR fluorescent secondary antibody: IRDye® 800CW Donkey anti-Rabbit IgG (H + L) [Li-cor, 1:5000 dilution]. Normalization was done by total amount of protein sample per lane, using the REVERT™ Total Protein Stain Kit [LI-COR, P/N 926-11010]. Immuno-stained bands were detected with the Odyssey NIR Western Blot Detection system and quantified by densitometric scanning using the ImageStudio Lite software.

### Lipid Peroxidation Assays

Ten thousand cells per well were seeded in a UV transparent flat-bottomed 96-well plate (Corning 3635). Lipid peroxidation (LPO) was measured using C11-BODIPY^581/591^ fluorescent dye, which shifts fluorescence emission when the polyunsaturated butadienyl portion of the molecule undergoes oxidation (Life technologies D3861). Cells were incubated for 30 min with 10 μM dye in PBS. Fluorescence was measured using a plate reader (SYNERGY H1) by simultaneous acquisition, samples were excited at 485 nm and emission was collected at 520/590 nm.

### Anti-Sense Oligonucleotide (ASO) Experiments

Custom designed 16-mer gapmer ASOs targeting the mutant *FTH1* mRNA (c.487_490_dup_gaat), were synthesized comprised of sugar-modified LNAs and phosphorothioate bonds in between nucleotides. LNA sugars were introduced at the 5’- and 3’-end of the oligo using a 3LNA-10-3LNA pattern. The sugar-modified LNAs (+) were used to increase binding affinity and nuclease stability of gapmers, while phosphorothioate (*) DNA was employed to elicit RNase H cleavage of the target^24^. LNA gapmer ASO, 5’+A*+A*+G*c*c*a*g*a*t*t*c*a*t*+T*+C*+G 3’ and scramble 5’ +G*+T*+A*T*a*t*c*c*a*g*a*t*a*+c*+G*+C 3’ were purchased from Integrated DNA Technologies (IDT). Cells were plated as described above for 24 h before transfection. Gymnotic LNA-ASO delivery was achieved using a 3 μM oligonucleotide concentration in complete cell culture medium. Cells were harvested for Western blot or RNA analysis 3 days after transfection. For lipid peroxidation studies, cells were harvested after 6 days of ASO treatment.

### Protein Modeling

FTH1 wildtype and FTH1 mutations were modeled in PyMOL based on the X-ray crystal structure of human heavy chain apo-ferritin (PDB ID: 5N27) including annotation of E-helix.

## Results

### Clinical Presentation*

*\*in compliance with medrxiv policy, exact ages, family relations, ethnic background and clinical presentation details were edited/removed as requested. Data available by contacting corresponding author*.

In brief, all five probands presented during early childhood with moderate to severe global developmental delay, with progressive spasticity, gait difficulties and ataxia. All probands were unrelated and from diverse ethnic backgrounds, including European, African American and South Asian. Imaging was consistent with pontocerebellar hypoplasia during infancy and later evolved to show evidence of mineralization suggestive of iron accumulation. Progressive cerebral volume loss was evident in patients that underwent serial imaging over time. Table 1 summarizes the salient clinical and neuroimaging features of the patient cohort.

**Table 1:** Clinical features of unrelated individuals with heterozygous *FTH1* variants*. *\*details edited/removed to comply with Medrxiv policies, ages provided in ranges*

| **Parameter** | **Pt 1** | **Pt 2** | **Pt 3** | **Pt 4** | **Pt 5** |
| --- | --- | --- | --- | --- | --- |
| Age at last evaluation, sex | 11-15yr, F | 16-20yr, F | 0-5yr, F | 0-5 yr, F | 11-15yr, F |
| **Neurologic Symptoms** |  |  |  |  |  |
| Dysphagia | no | yes | yes | yes | Yes |
| Involuntary movements | mild dystonic posturing | dystonia, athetosis | yes | startle | Startle |
| Epilepsy | yes | single seizure | yes | no | No |
| EEG | multifocal sharp waves, more frequent right posterior | generalized background slowing | right posterior sharps, myoclonic seizures | abnormal background without interictal discharges during sleep/wake | No epileptiform activity |
| **Neuroimaging** | abnormal | abnormal | abnormal, PCH | abnormal | Abnormal |
| White matter | hyper T2 periventricular | decreased | decreased | decreased | Supratentorial white matter volume loss |
| Cerebellum | progressive atrophy | hypoplasia, progressive atrophy | hypoplasia | hypoplasia | atrophy |
| MRI SWI+ signal abnormalities | yes | yes | no | yes, not intraparenchymal (likely due to birth) |  |
| Basal ganglia abnormal | yes | yes | yes | yes | yes |
| **Genetics/Diagnostics** |  |  |  |  |  |
| Metabolic testing | hi lactate/pyruvate ratio; nl pl aa, acp, uoa | minimal lactate, Wilson nl |  |  | Nl uoa, acp |
| Other genetic tests | SNP 7q31.1 del VUS | Rett, Angelman, Fragile X nl | SNP normal | SNP 8p12 dup VUS, 12p12.1 del VUS | SNP normal |
| **Whole exome** |  |  |  |  |  |
| *FTH1* variant type | Truncation | Truncation | Truncation | Truncation | Truncation |
| cDNA, protein position | c.487_490 dupGAAT, p.Ser164*, de novo | c.512_513delTT, p.Phe171*, de novo | c.512_513delTT, p.Phe171*, not mat | c.512_513delTT, p.Phe171*, de novo | c.512_513delTT p.Phe171*, de novo |
| **Systemic/Labs** |  |  |  |  |  |
| Ophthalmologic | normal (9yr) | abnormal | nd | abnormal | Cortical visual impairment |
| CV/Renal involvement | no | no | no | no | No |
| GI/constipation | N/A | yes | yes | yes | Enteral feeds |
| LFTs | normal | normal | normal | N/A |  |
| **Iron Studies** |  |  |  |  |  |
| Ferritin, ng/mL (reference) | 7.9 (10-150) | 6 (12-114) | 10.6 (13-150) | 50.7 (5.3-99.9) | 162 (6-67) |
| Serum iron, ug/dL | 28 (28-147) | 35 (29-189) | 47 (33-151) | 15.5 (4.8-25.4 umol/L) | 111 (27-164) |
| TIBC, ug/dL (reference) | 391 (250-450) | nd | 588 (250-425) | nd | 252 (271-448) |
| Transferrin saturation (%) | 11 (20-50) | 9 (10-47) | 8 (15-55) | nd | 44 (15-45) |
| Hemoglobin, g/dL | 10.3 | 11.3 | 10 | 12.6 | 14.1 |

### Exome Results

Exome sequencing was performed. Heterozygous *FTH1* variants were found in all five unrelated probands, with four probands sharing a recurrent variant. Additional rare variants were also considered as candidates but were not shared among the affected individuals. The *de novo* nature of the *FTH1* variants was confirmed in all four probands where trio analysis could be performed. Proband 1 had a novel heterozygous, *de novo* variant c.487_490dupGAAT (p.S164*) in *FTH1*, reported by the clinical laboratory as a candidate. She was also found to carry a paternally inherited variant of unknown significance (p.R253W) in the *TOE1* gene, which is associated with a recessive form of pontocerebellar hypoplasia (PCH7). The copy number variant of unknown significance (*IMMP2L* deletion) was found to be inherited from an asymptomatic parent. Patient 2 had a novel heterozygous, *de novo, FTH1* variant c.512_513delTT, (p.F171*), found using clinical exome sequencing, identifying a candidate gene. Two other *de novo* gene variants were found with low quality and there was no human disease correlate. Probands 3, 4, and 5 were found to have the same recurrent *FTH1* variant as proband 2, c.512_513delTT (p.F171*), although all these individuals were unrelated. The variant was confirmed as *de novo* in probands 4 and 5 as well. Proband 3 underwent a duo exome, as a paternal sample was not available; the variant was not found in the maternal sample. Given the c.512_513delTT, (p.F171*) variant was found in four out of five affected individuals in the cohort, the variant was suspected to be the top candidate. *FTH1* has four exons, and all variants in the probands occurred in the last exon.

The recurrent dinucleotide deletion c.512_513delTT, leading to p.F171*, was not present in gnomAD v.2.1.1. or v.3.1.2., nor was the dup variant c.487_490dupGAAT leading to p.S164*. Genic intolerance to variation in the population data also suggested the gene’s potential importance. Both variants were in the final exon of the *FTH1* coding sequence suggesting likely escape from NMD. In addition, occurring in the final exon of *FTH1*, the variants ae predicted to lead to a truncated C-terminus (∽12-20 amino acids) in the FTH1 protein.

### Distinct Neuroimaging Overlapping with PCH and NBIA syndromes

Several distinct neuroimaging findings were identified in the probands with stopgain variants in the final exon of *FTH1*. Brain imaging of proband 1 (Figure 1A-D) showed marked areas of abnormal iron deposition along with gliotic changes involving the anterior segment of the globus pallidi, giving the so-called eye of the tiger sign (Figure 1C). Moreover, additional findings including progressive volume loss involving the cerebellum, particularly the vermis, and interval appearance of volume loss and signal changes in the supratentorial white matter were noted over time when serial imaging was reviewed (baseline imaging shown Figure 1D). Similarly, serial imaging of proband 2 also demonstrated progressive volume loss, persistent basal ganglia signal abnormalities and cerebellar hypoplasia. Imaging for proband 3 was remarkable for areas of abnormal iron mineral deposition that were noted in the basal ganglia as early as the first MRI performed before 1 year of age. The signal changes were not selective to any structure in the basal ganglia and were better observed in the gradient echo, and magnetization transfer T1 sequences. Noteworthy additional imaging features are shown from MRI around four years later, including severe pontocerebellar hypoplasia with severe atrophy of the midbrain in the axial plane giving the “figure 8” appearance (Figure 1H), along with T2 crossed linear hyperintensities, representing selective degeneration of transverse pontocerebellar tracts and median pontine raphe nuclei (“hot cross bun sign”) (Figure 1I). The “hot cross” bun sign represents a classic imaging pattern related to other neurodegenerative disorders, particularly the cerebellar subtype of multiple system atrophy, and different types of spinocerebellar ataxia^25^. The association between pontocerebellar hypoplasia and imaging features of axonal tract degeneration (hot-cross bun sign) involving the pons suggest that this disease has a progressive course likely with a prenatal onset. Additional findings including severe white matter volume loss particularly involving the frontal, parietal, and temporal lobes along with diffusely thin corpus callosum were also noted (Figure 1E-G).

**Figure 1.**
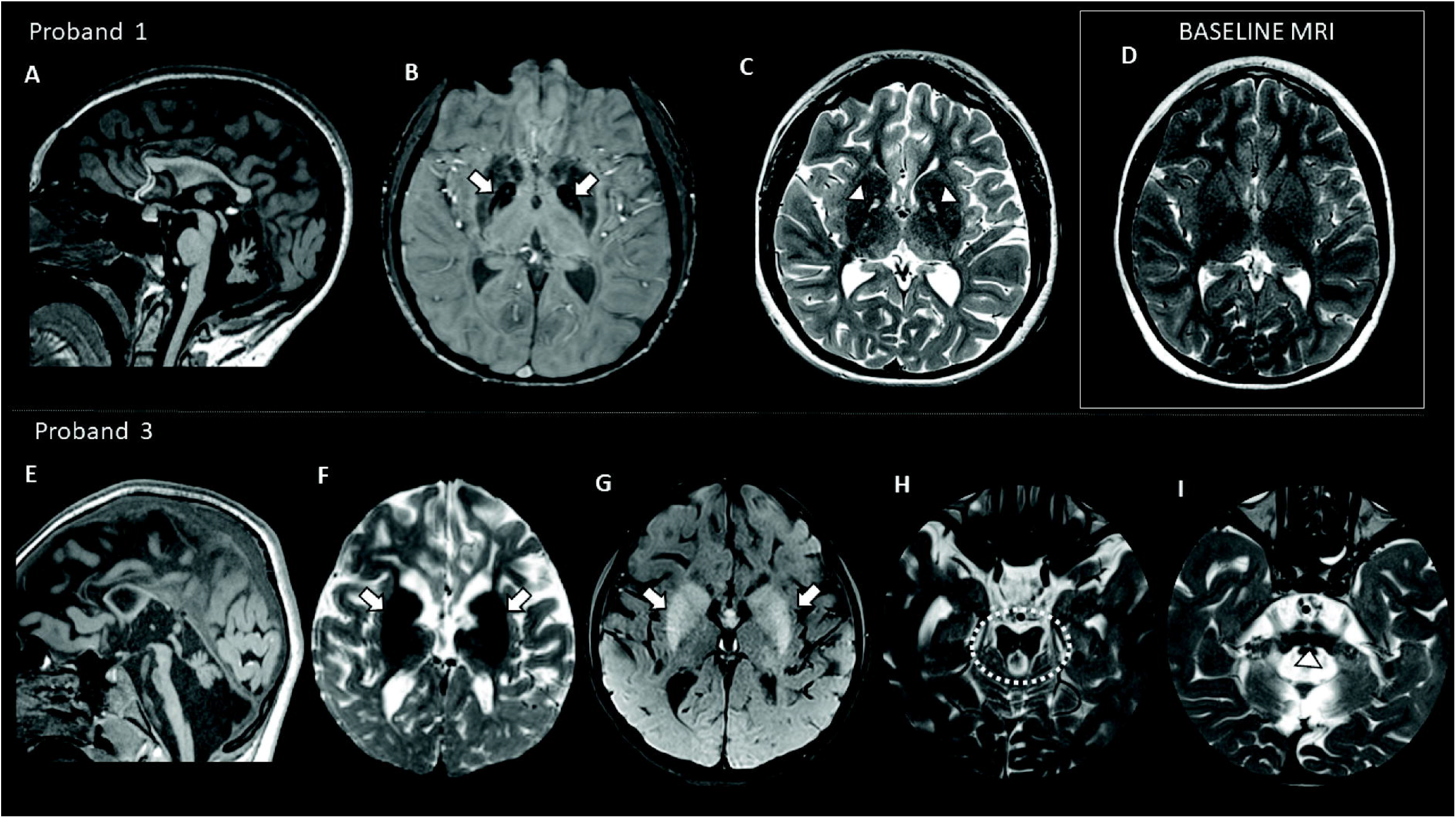
Neuroimaging findings of individuals with heterozygous *FTH1* variants. Proband 1: 11-15 years old female brain MRI (A-C), and baseline brain MRI when she was 6-10 years old (D). (A) Sagittal T1WI shows severe hypoplasia and atrophy of the vermis along with a diffusely thin corpus callosum. (B) Axial SWI shows artifacts within the globus pallidi corresponding to abnormal areas of iron deposition (arrow, B). (C) Axial T2WI demonstrate selective hyperintense signal involving the anterior segment of the globus pallidi, giving the eye of tiger sign (arrowhead, C), feature not observed in the baseline MRI axial T2WI, 5 years before (D). Proband 3: 0-5 years old female brain MRI (E-I) (E) Sagittal T1WI shows severe pontocerebellar hypoplasia along with a diffusely thin corpus callosum. Axials gradient echo, and magnetization transfer T1 show dark artifacts (F) and T1 shortening (G) within the basal ganglia corresponding to abnormal areas of mineralization (arrows, F, and G) along with severe white matter volume loss. Severe atrophy of the midbrain giving a “figure 8” appearance of this structure (dotted circle, H) and T2 crossed linear hyperintensities (arrowhead I), representing the hot-cross bun sign.

Selective cerebellar atrophy, particularly involving the vermis, with or without the involvement of the brainstem, along with progressive white matter volume loss and evidence of iron deposition in the basal ganglia in the early stages of the disease represent the most common imaging findings in the patients imaged with *FTH1* variants. Overall, for cases reviewed, the supratentorial atrophy appears mainly related to white matter volume loss. Moreover, additional imaging markers including the eye of the tiger sign in the globus pallidi and the hot cross bun sign in the pons were noted in some of the patients, evincing that these findings are not necessarily pathognomonic for PKAN and multiple system atrophy type C, respectively.

### Pathological Findings

Neuropathological studies were performed for Proband 2, who died of complications of the disease in her 20s. She experienced dysphagia, weight loss, menorrhagia, and lethargy, and despite transfusion, hospitalization and stabilization, she continued to decline and signs of pneumonia were found on autopsy. There was diffuse and extensive involvement of brain, brainstem, and spinal cord with striking involvement, and patchy destruction, of the basal ganglia and midbrain, including the red nucleus and substantia nigra. Eosinophilic intracytoplasmic and intranuclear inclusions were present diffusely (Figure 2A), including NeuN positive neurons (Figure 2B). Extracellular deposits were particularly prominent in the white matter and basal ganglia, where Prussian blue staining revealed patchy iron deposition (Figure 2C). Inclusions were also found in the cortex and cerebellum. The eosinophilic inclusions and deposits were diffusely immunopositive for FTH1 (Figure 2D). Pathological analyses revealed rare iron staining in the gastrointestinal system, suggesting other cells outside of the nervous system may be subtly affected.

**Figure 2.**
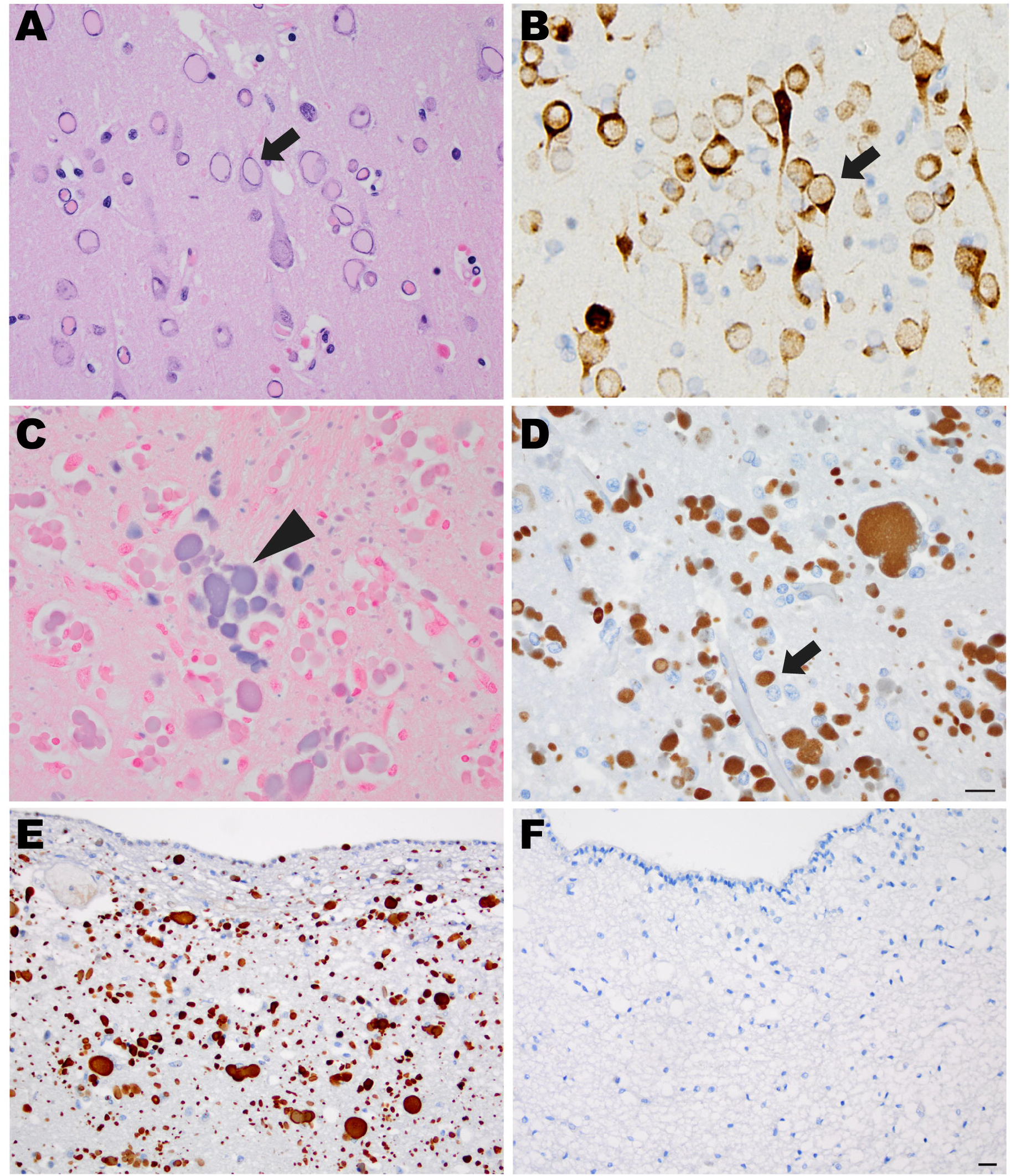
Neuropathology and ferritin staining. Representative H&E images from (A) frontal cortex show intracellular inclusions (arrow) (B) NeuN immunohistochemistry showing intracellular inclusions present in neurons (arrow). There was patchy iron deposition shown in the basal ganglia by Prussian blue stain (arrowhead) (C), and the inclusions immunostained for ferritin heavy chain protein FTH1 (arrow) (D,E). Negative control: unaffected brain (F). Magnification is 400x (A-D) and 200x (E-F), scale bar is 20 μm.

### *FTH1* Transcripts and Increase In Cellular Ferritin Protein Levels

As all variants identified in the patients were nonsense variants predicted to lead to premature protein truncation^26^. Truncating variants in the last exon of a gene are typically expected to escape nonsense mediated decay (NMD). We first assayed total *FTH1* mRNA and protein levels in patient fibroblasts compared to control lines. Both heavy (Figure 3A) and light chain (Figure 3B) ferritin protein levels were elevated in patient fibroblasts relative to controls. Therefore, *FTH1*-variant cells demonstrate an increase in ferritin heavy chain protein levels, which suggests the variant mRNA transcript does not lead to haploinsufficiency. To determine how *FTH1* variants affected total *FTH1* transcript levels, we assayed mRNA levels by RT-PCR. *FTH1* mRNA expression levels were reduced in patient cells vs controls, by around 20 percent (P1=78% of control, P2=83%, Figure 3C). *FTL* mRNA transcript levels were either lower or unchanged in patient cells, only statistically significant in the P2 line. The results suggest other mechanisms influencing ferritin protein levels beyond mRNA translation, which could include posttranslational regulatory mechanisms and/or effects of the variants in protein stability or regulation^27^.

**Figure 3.**
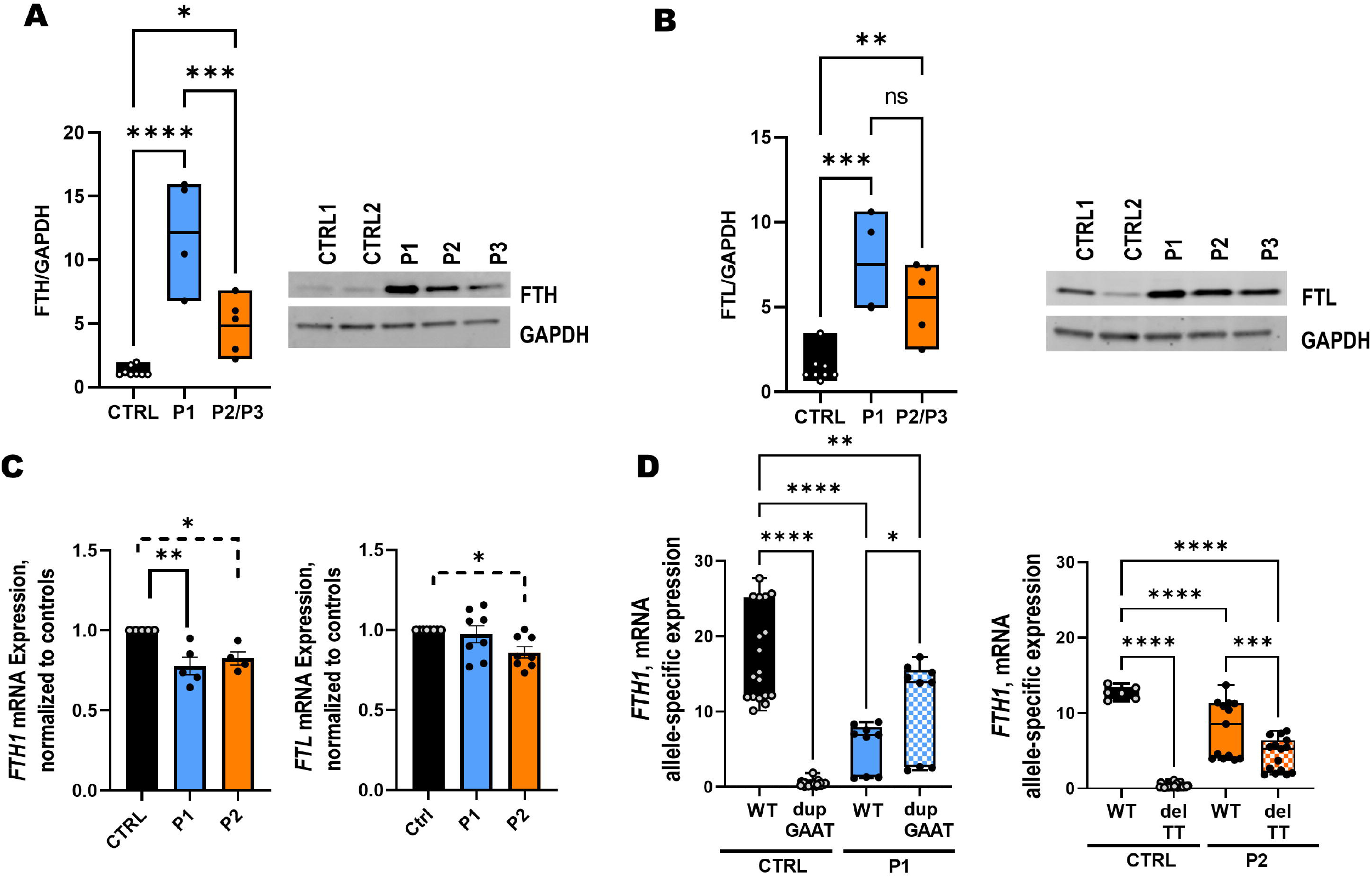
*FTH1 de novo* variant effects on ferritin heavy and light chain protein and mRNA levels. A-B) Quantification of protein levels and representative immunoblots showing FTH and FTL protein levels in primary patient-derived fibroblasts (filled circles) vs controls (open circles). All *FTH1*-variant cells (shown as P1, P2, P3) showed elevated levels of both light and heavy chains of ferritin relative to controls. Quantification was analyzed by genotype using two independent cell lines per genotype, except P1 which is the only available line with the p.S164* variant [P1= *FTH1* c.487_490 dupGAAT (p.S164*); P2 and P3= *FTH1* c.512_513delTT, (p.F171*), Controls=FTHctrl and Coriell 8400 lines]. C) Quantification of total *FTL and FTH1* mRNA transcripts by RT-PCR in patient fibroblasts relative to controls (mean levels relative to control=1: P1 0.778±0.05, P2 0.825±0.06). D) RT-PCR for allele-specific *FTH1* mRNA transcripts, performed for each genotype. *FTH1* mutant transcripts are detectable with mutation-specific primers in patient fibroblasts. In P1, mutant transcript levels were higher that wild type transcripts, while in P2, wild type levels were higher than mutant transcript. In both genotypes, however, the data demonstrate that nonsense mutant transcripts were present and escape nonsense mediated decay. All data represented as mean ± SEM; Data analyzed with one way Anova with Tukey’s multiple comparisons test; *p<0.05, **p<0.01, ***p<0.001, ****p<0.0001

To further determine whether the mutant transcripts are able to escape nonsense mediated decay, we designed mutant specific primers for each variant (c.487_490dupGAAT for P1 and c.512_513delTT for P2). We quantified wild type vs mutant transcripts simultaneously by RT-PCR. Mutant-specific mRNA transcripts were readily detectable in patient-derived cells and not in controls (Figure 3D) demonstrating escape from NMD. Mutant mRNA transcripts were not only clearly detectable, but in the case of P1, mutant transcript levels were higher than those of the wild type allele.

In order to evaluate the subcellular localization of ferritin heavy chain, *FTH1* patient fibroblasts were immunostained using anti-ferritin light and heavy chain antibodies, as well as the lysosomal marker LAMP1. Ferritin can be degraded in the lysosome, also known as ferritinophagy. Fibroblasts showed ferritin signal in the expected predominant cytoplasmic distribution, with aggregate-like structures visible in patient cells (Figure 4C) and not in control (Figure 4A). Iron exposure with ferric ammonium citrate (FAC) for 3 days led to expected increase in ferritin signal in both control (Figure 4B) and patient cells (Figure 4D); ferritin staining and aggregate-like structures were more robust in patient cells. We did not observe significant colocalization of heavy nor light chain ferritin in lysosomes in fibroblasts (Figure 4A-D).

**Figure 4:**
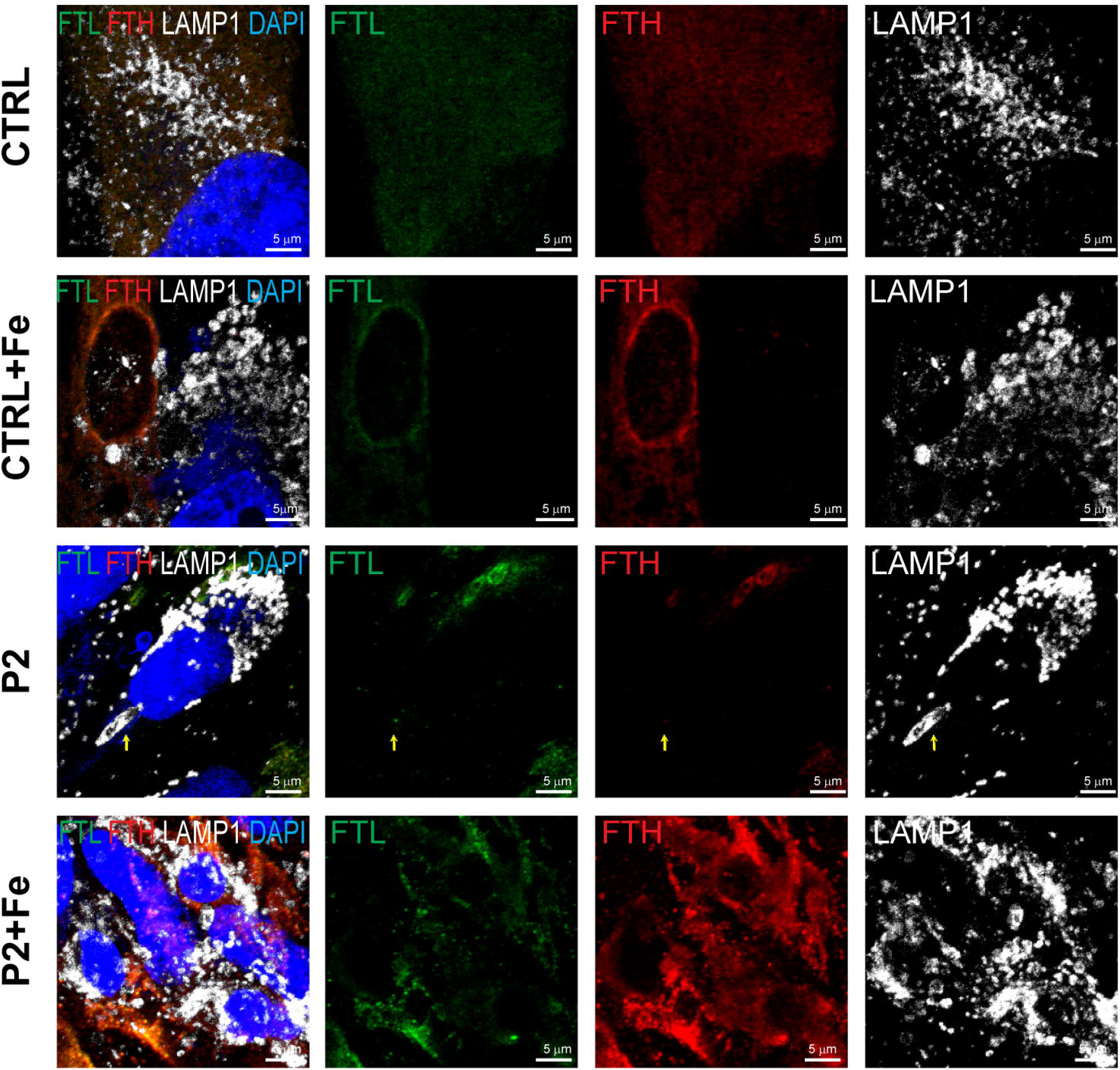
Immunocytochemistry for ferritin subcellular localization (A-D). Cells were stained with anti-FTL (green), anti-FTH (red), lysosomal LAMP1 (white), and nuclear stain DAPI (blue). Scale bar = 5 μm. For exogenous iron treatment (+Fe), cells were treated with 150 μg/mL FAC for 3 days.

### Cellular Iron Content and Susceptibility to Oxidative Stress

Elemental iron content was measured by the highly sensitive technique of inductively coupled plasma optical emission spectroscopy (ICP-OES), at baseline and after iron treatment (FAC, ferric acid citrate 150 μg/mL). No significant differences in iron content were detected in patient fibroblasts at baseline, when tested up to 7 days in culture (Figure 5A, untreated controls shown as lower dark grey bars). With iron exposure, as expected, all cell lines showed statistically significant increases in iron content (by 3, 7 days) relative to respective untreated controls (top bars). Nevertheless, there was no significant difference in iron content in control vs patient fibroblasts assayed at days 1 or 3 of treatment. At 7 days of iron exposure, the p.S164* variant cells showed a significant increase in iron content relative to controls, but there was no significant difference between the two p.F171* cell lines and controls, suggesting there may be variability in the cellular effects of the ferritin variants.

**Figure 5.**
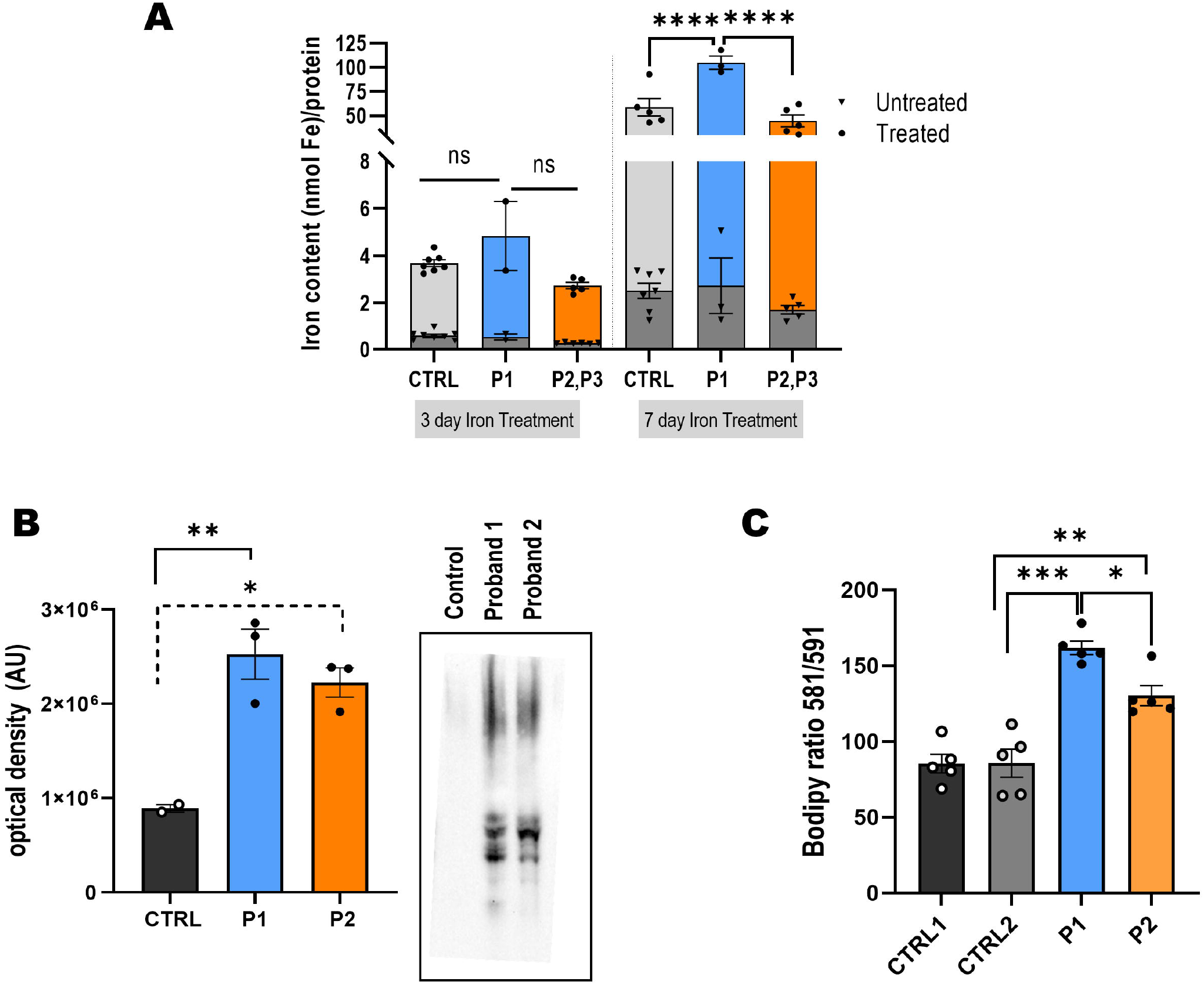
Iron Content and Oxidative Stress Markers. A) Iron exposure experiments in patient-derived cells and iron quantification. Fibroblasts were treated with FAC, 150 μg/mL for 3 or 7 days and assayed with inductively coupled plasma optical emission spectroscopy (ICP-OES) as described. Dark grey base bars are untreated controls for each cell line. B) Oxidative stress assay: oxyblot immunodetection of carbonyl groups, quantified by densitometry in control vs patient cells. C) Lipid peroxidation levels were assayed with C11-BODIPY ^581/591^ and quantified by plate reader. For most assays n≥2 cell lines per genotype (except for P1 as only one line available); for each cell line assays with technical triplicates. All data represent mean ± SEM. *p<0.05, **p<0.01, ***p<0.001.

Because cellular iron dysregulation can be associated with oxidative stress and is hypothesized to be relevant to neurodegenerative processes^19,28^, we tested whether the *FTH1* cells showed signs of oxidative damage. Because increased oxidative stress is known to cause protein and lipid peroxidation, we assayed for such markers in patient cells. Oxyblot assays for protein oxidation were performed (Figure 5B), showing an increase in total oxidized proteins in P1 and P2 fibroblasts. To assess lipid peroxidation (LPO)^3,29^, the lipid peroxidation sensor B11-BODIPY was used. *FTH1*-mutant cells showed higher levels of peroxidized lipids compared to controls (Figure 5C), suggesting *FTH1*-variant cells demonstrate intrinsic differences in oxidative stress.

### Structural Modeling Suggests *FTH1* Variants Alter Ferritin’s Pore

To model the potential effects of *de novo* heterozygous *FTH1* variants on the ferritin 24mer complex, we analyzed 3-dimensional structures using PyMOL and the X-ray structure for heavy ferritin chain (PDB ID: 5N27). The wild-type FTH subunit is comprised of a four α-helix bundle (helices A-D) connected via a flexible loop to a shorter E-helix. The 3-dimensional protein structure changes with the p.S164* mutation, which deletes the E-helix, while the p.F171* mutation leads to truncation of the E-helix. Alteration of the E-helix results in malformation of the 4-fold symmetric pores, which are important for iron retention^30-32^. We predict from structural analysis that the pore formed by p.S164* should be substantially larger than wild-type. This structural defect is exacerbated when all four subunits that comprise the pore have a modified E-helix (Figure 6).

**Figure 6:**
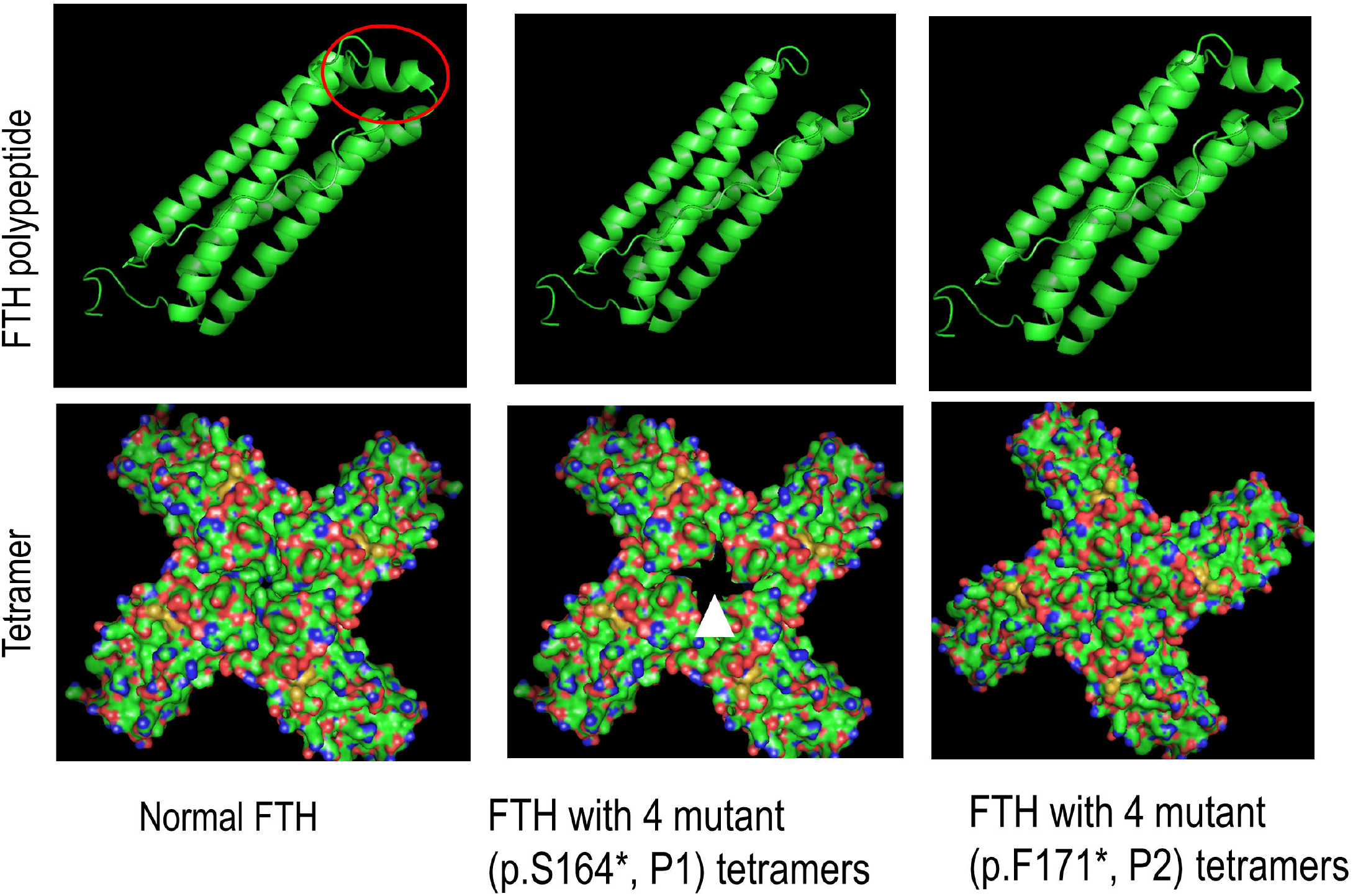
Structural modeling of *FTH1 de novo* variants predict truncation of E-helix (circle) and are predicted to affect ferritin’s pore size (arrow head) for p.S164* variant and pore depth (not evident in the schematic, but the depth of the pore is predicted to be 4.5 Å shallower than wild-type due to the missing terminal residues) for p.F171* variant.

Interestingly, assembly of the p.F171* subunits is predicted to minimally impact pore size at the four-fold axes. Nevertheless, for the p.F171* variant, the depth of the pore is predicted to be 4.5 Å shallower than wild-type due to the missing terminal residues. The p.F171* variant is also missing K173, and this positive charge is potentially important for preventing iron from leaving the ferritin cavity through the pore. These changes could lead to altered pore function, ferritin structure or reduced ferritin iron binding capacity.

### Allele-Specific Oligonucleotide (ASO) Rescues *FTH1* p.S164* Variant Cells

Based on our findings of elevated ferritin protein levels in patient-derived cells and RT-PCR revealing that mutant *FTH1* alleles are detectable in patient but not control cells, we hypothesized that *FTH1* variants exert a dominant negative effect by escaping NMD. Therefore, we used the allele-specific mRNA assay to test the effectiveness of an ASO designed to suppress the expression of the mutant allele in P1 cells. We selected the P1 cells as the structural (larger effect on the pore) and experimental data (significant iron accumulation upon exposure) suggested a more robust cellular phenotype for the P1 variant (p.S164*) compared to the recurrent variant (p.F171*). We found that mutant allele mRNA expression was suppressed with ASO treatment, but not with a scrambled control (Figure 7A). By suppressing the mutant allele in the P1 line, we could also detect diminished increase in lipid peroxidation (LPO) to control levels (Figure 7B). Furthermore, treatment with mutant-specific ASO partially rescued the elevated ferritin heavy chain protein levels in P1 cells to near control levels (Figure 7C).

**Figure 7.**
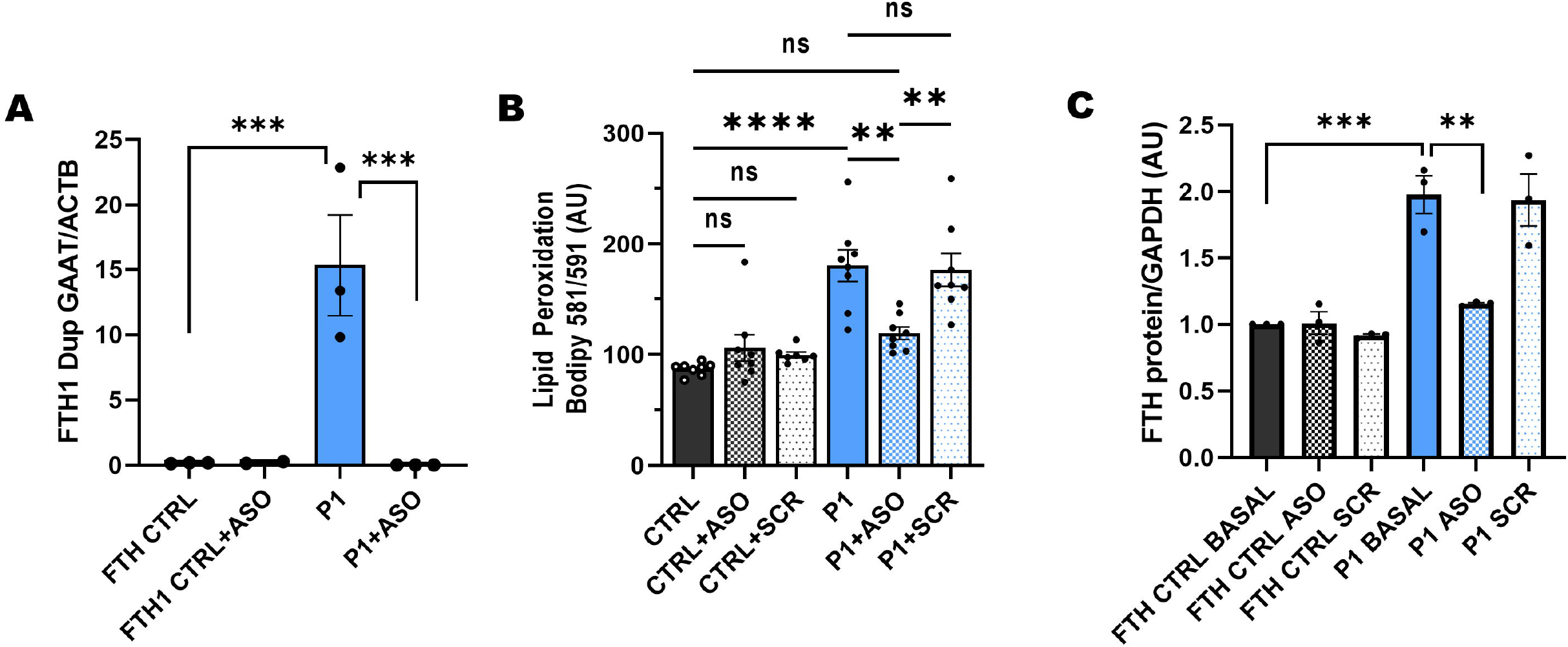
Antisense oligonucleotides (ASO) directed toward *FTH1* variant in Proband 1 (c.487_490 dupGAAT, p.Ser164*) in fibroblasts. (A) RT-PCR was used to detect the mutant allele specifically, verifying its detection in patient fibroblasts but not in control. The P1 targeted ASO specifically suppressed expression of the mutant allele and not wild type. (B) Treating P1 cells with the ASO restored lipid peroxidation levels, while scrambled (SCR) control had no effect (n=3 experiments with ≤<u>4 technical replicates per group).</u> (C) ASO reduces ferritin heavy chain protein to near control levels. All data represented as mean ± SEM; Data analyzed with one way Anova with Tukey’s multiple comparisons test; *p<0.05, **p<0.01, ***p<0.001, ****p<0.0001

## Discussion

This report documents a novel neurodevelopmental disorder associated with heterozygous *de novo FTH1* variants, with symptoms, neuroimaging and histological findings supporting a novel neuroferritinopathy. Probands presented in early childhood with neurodevelopmental delay and some later developed progressive neurological symptoms including ataxia, seizures, and spasticity. Notably, early brain imaging findings exhibited overlap with some forms of congenital pontocerebellar hypoplasia, but signal abnormalities suggesting iron accumulation became evident later in life. Our data suggest that the described *FTH1* variants exert a dominant negative effect, perhaps due to impaired ability of ferritin to store iron as the truncated E-Helix may alter ferritin’s pore size (p.S164*) or depth (p.F171*). This appears to result in oxidative stress, as evidenced in functional validation studies in patient-derived fibroblasts. This novel disorder is further distinguished by specific neuroimaging features, including the eye of the tiger sign, classically described in the context of PKAN^33^, some cases with evidence of selective degeneration of the transverse fibers of the pons and median pontine raphe nuclei, leading to the “hot cross bun” sign, and severe atrophy of the midbrain in the axial plane giving the “figure 8” appearance along with severe leukomalacia, classically seen in *AMPD2*-associated PCH type 9^34^. These data expand the differential diagnosis of these neuroimaging signs to include *FTH1*-associated neuroferritinopathy.

The individuals described in this study presented with neurodevelopmental delays early in life, whereas in *FTL*-associated neuroferritinopathy, symptom onset typically occurs in adulthood, first with movement disorders (mean age of onset 39 years), followed by cognitive decline^17,18^. In addition to endogenous production in the brain, cellular systems and animal models have shown that heavy chain ferritin protein (but not L ferritin) is able to cross the blood-brain barrier^35,36^. The possibility that the brain may accumulate FTH from the body could potentially contribute to the childhood symptom onset in patients with *FTH1*-related disease compared to patients who present at middle age *FTL*-related disease. Additional studies would be needed to test these possibilities.

Despite different ages of onset, both disorders share imaging features such as iron accumulation and clinical evidence of neurodegeneration. The neuropathological findings in our probands with *FTH1* variants are reminiscent of those described in *FTL*-associated neuroferritinopathy, including immunohistochemical staining showing ferritin positive aggregates and iron accumulation in the brain^37^. *FTH1*-associated neuroferritinopathy appears to have early pontocerebellar involvement as a distinct feature. Disease models of *FTL*-associated neuroferritinopathy have also been shown to exhibit ferritin aggregates, oxidative stress and higher lipid peroxidation levels^28,38,39^. We found that upon iron exposure, patient-derived cells showed significantly increased levels relative to controls. A potential limitation is that fibroblasts may not completely recapitulate the effects of *FTH1* variants in other cell types, particularly neurons. In addition, the different ratios of ferritin heavy and light chains in different cells could also be important. Future studies in animal and/or IPS models should examine the tissue specific-sequalae of truncating the E-helix in the central nervous system.

Similar to adults with *FTL*-associated neuroferritinopathy, our cases do not show significant systemic iron overload or overt hyperferritinemia. Serum ferritin levels were low or normal in all but one proband (Table 1). Notably, 5’ non-coding variants in both *FTL* and *FTH1* have been previously associated with iron overload. In hyperferritinemia with or without cataracts (MIM 600866)^15,40^, heterozygous variants occur in the 5-prime non-coding *FTL* region that binds to iron regulatory elements^41^. It is thought that these variants result in disrupted regulation of ferritin light chain translation, leading to hyperferritinemia. In hemochromatosis type 5 (MIM 615517), a single family was reported with missense variant in the 5’-UTR of *FTH1* (p.A94U) altering affinity for IRP (iron regulatory protein) binding^42^. On the other hand, pathogenic variants leading to neuroferritinopathy in *FTH1* and *FTL* both show recurrent C-terminus nucleotide deletions/duplications leading to frameshift, truncating the E-Helix of the ferritin protein subunits^3,38^. The data here supports dominant negative action of mutant ferritin, with structural modeling data suggesting alterations in ferritin pore structure. None of the five patients had signs of hemochromatosis. However, some patients in our cohort had low serum ferritin levels (Table 1), which may be consistent with serum ferritin findings in *FTL*-neuroferritinopathy^13^.

Structural studies suggest that the ferritin protein E-helix is fundamental for the stability and assembly of the protein complex. Although this domain does not have a ferroxidase function nor contain an iron binding site, it can be flipped in or out of the nanocage^43^, thereby affecting its capacity for binding and mineralizing iron. Previous studies with FTL confirmed that amino acid deletions in the FTL C-terminus resulted in iron-induced precipitation and revealed iron mishandling^30^. FTL E-helix mutations have been linked to the formation of ferritin inclusion bodies, which highlights the significance of the wild-type E-helix in producing fully functional ferritin protein. Targeted mutagenesis studies with murine ferritin heavy chain protein have shown that truncating the E-helix contribute to poor ferritin stability and solubility and poor sequestration of cellular iron^44^. In alignment with these previous data, the structural *in silico* data presented here predicts that C-terminal truncating variants in *FTH1* alter the E-helix, affecting ferritin pore size and/or depth, which would be expected to alter ferritin’s iron storage capacity.

The data reported herein, including elevated cellular ferritin levels, detection of mutant mRNA despite nonsense variants, and the potential rescue of cellular phenotypes with targeted mutant allele knockdown with ASO, suggest that the pathogenic mechanism of the *FTH1* heterozygous variants is a dominant negative effect, as opposed to haploinsufficiency. Concordant with this, previous mouse studies demonstrate that haploinsufficiency of *Fth1* does not result in a clear phenotype^45^. Suppressing mutant allele expression with ASO provides proof of principle for a strategy to possibly ameliorate pathogenic *FTH1* expression in cells.

In summary, the results presented here support a new human disease association for the iron homeostatic gene, *FTH1*. Heterozygous *de novo* stopgain variants in the final exon of *FTH1* are associated with a novel pediatric neuroferritinopathy within the spectrum of NBIA disorders. Our findings suggest disease-causing *FTH1* variants lead to truncation of the E-helix of ferritin heavy chain, which alters protein function by altering the ferritin pore size and/or depth. Neuropathology confirms ferritin aggregates in the brain and widespread evidence of neurodegeneration. The data are consistent with the proposed disease mechanisms for *FTL*-associated hereditary neuroferritinopathy, suggesting some degree of shared downstream consequences, regardless of whether mutations occur in the ferritin light or heavy chain. Finally, the cellular effects of pathogenic *FTH1* variants can be modified with antisense oligonucleotides, raising the possibility that similar therapeutic approaches may be applicable for patients with neuroferritinopathy regardless of genetic etiology.

## Data Availability

All data produced in the present study are available upon reasonable request to the authors.

## Acknowledgements

We wish to thank the patients and their families who participated in this study and also acknowledge their philanthropic support via the NBIA Fund at the Children’s Hospital of Philadelphia. JTS thanks the Marcus Program in Precision Medicine and the Benioff Children’s Hospital, University of California San Francisco for their funding and support. IJD thanks the NSF, CHE-1905203. XOG thanks the Robert Wood Johnson Foundation (Harold Amos Faculty Development Program), NINDS (1K02NS112456-01A1) and the Burroughs Wellcome Fund (CAMS Award). Also, CHOP’s Roberts Collaborative for Genetics and Individualized Medicine (to XOG). We acknowledge the UCSF Brain Tumor SPORE Biorepository (NIH/NCI P50CA097257 JJP) for providing histology services. We thank UCLA Genomics Center, GeneDx and GeneMatcher^46^.

## Author Contributions

JTS and XOG contributed to the conception and design of the study; JTS, JATH, CNM MPP, MFH, AS, LD, ND, RML, CB, JCL, JJP, CAPFA, IJD, XOG contributed to the acquisition and analysis of data; JTS, JATH, CNM, AS, MFH, CAPFA, XOG contributed to the drafting a significant portion of the manuscript or figures. All authors contributed to copy editing and approval.

## Declaration of Interests

The authors declare no competing interests.

## Data and Code Availability

The data described during this study will be made available upon reasonable request.

## Social Media Summary

The report describes *de novo* variants in *FTH1*, encoding ferritin heavy chain are linked to a novel pediatric disease in the spectrum of NBIA (neurodegeneration with brain iron accumulation). Our data suggest that C-terminus variants truncating the E-Helix of the ferritin heavy chain lead to brain ferritin accumulation and cellular oxidative stress, which can be ameliorated by antisense oligonucleotides. Twitter Handles: @drxilma, @chayasays

## Notes

### Competing Interest Statement

The authors have declared no competing interest.

### Funding Statement

This study was made possible by philanthropic support via the NBIA Fund at the Childrens Hospital of Philadelphia. JTS thanks the Marcus Program in Precision Medicine and the Benioff Childrens Hospital, University of California San Francisco for their funding and support. IJD thanks the NSF, CHE-1905203. XOG thanks the Robert Wood Johnson Foundation (Harold Amos Faculty Development Program), NINDS (1K02NS112456-01A1) and the Burroughs Wellcome Fund (CAMS Award). Also, CHOP Roberts Collaborative for Genetics and Individualized Medicine (to XOG). We acknowledge the UCSF Brain Tumor SPORE Biorepository (NIH/NCI P50CA097257 JJP) for providing histology services.

### Author Declarations

IRB of The Children's Hospital of Philadelphia gave ethical approval for this work. IRB of the University of California San Francisco gave ethical approval for this work. IRB of the Baylor College of Medicine gave ethical approval for this work. IRB of the Hospital for Sick Children gave ethical approval for this work.

